# Estimation of soleus muscle activation patterns from lower limb kinematics during normal level walking using a deep neural network model

**DOI:** 10.1101/2025.10.14.25337892

**Authors:** Jobelle B. Hernandez, Oliver Gu, Aymen Elassa, Mariam Sharobim, Samira Santana, Jongsang Son

## Abstract

The purpose of this study is to develop a deep neural network model for predicting soleus muscle activation patterns from time-series lower-limb joint angles collected during level ground walking at different speeds and to evaluate the prediction performance. An open dataset was used to obtain full lower-limb kinematics, including pelvis, hip, knee, and ankle joints, and soleus muscle activation patterns from 20 control adults. Long short-term memory (LSTM)-based deep learning models were developed and then evaluated for prediction performance by conducting both the random split cross-validation (CVM1) and the leave-one-subject-out cross-validation (CVM2). For both cross-validation methods, the developed models yielded promising error and regression metrics such as the root mean square error and coefficient of determination. However, the CVM2 demonstrated that the prediction performance can be sensitive to individual datasets. The subject factors, such as age, sex, and walking speed, appear to have a negligible effect on the prediction performance for the CVM1. This study demonstrated the feasibility of the developed models to be a template for a potential tool that quantifies muscle activation patterns from joint angles during level ground walking at different speeds.

## Introduction

A quantitative description of gait can provide insights into understanding potential underlying mechanisms of an abnormal walking pattern especially in an individual with musculoskeletal, cardiovascular, or neurologic conditions and planning the most appropriate and effective treatment plans [1–4]. In this aspect, the analysis of muscle activation patterns during walking is one of the most important, useful data in clinics [5]. However, gait analysis in clinical settings is typically qualitative as clinicians likely rely on observation of patients and questionaries/surveys during in-person examinations. Surface electromyography (sEMG) is widely used to record electrical activities associated with muscle activation and contraction (albeit mostly in research settings), thus providing physicians with additional information to better understand abnormal gait patterns and potential causes of such gait abnormality [6]. Such additional information would lessen the influence of the subjectivity of qualitative analysis. This highlights the clinical relevance of gait quantification and assessment through muscle activation.

Although various methods have been introduced for quantitative gait analysis, the majority is limited to research settings and seldom translated into routine clinical practice [5]. Quantitative analyses involving optical motion capture and EMG measurement systems are considered the gold standard. Despite the reliability and accuracy of gold standard methods, their complicated set-up and operation, high costs, and lack of globally accepted standards limit their feasible application in clinical settings [5, 7]. These drawbacks appear to outweigh the benefits of its integration into clinical practice. Although there are several potential alternative options that can be used in muscle activation analysis, such as mechanomyography (MMG) [8], sonomyography (SMG) [9, 10], and optomyography (OMG) [11], these methods have not established a large volume of data in muscle activation patterns especially during dynamic movements. Likely due to these limitations, quantitative motion analysis has not been extensively adopted in routine clinical practice, limiting the ability of clinicians to interactively utilize the current level of function of a patient to plan the personalized rehabilitation protocols and to track the patients’ function during rehabilitation sessions.

More recently, machine-/deep-learning approaches accompanied by wearable sensor technology have been considered a promising alternative to conduct quantitative human movement analysis. For instance, inertial measurement units (IMUs) have been used as input data to machine-/deep-learning models designed in estimating not only kinematic variables such as joint angles [12–16] but also kinetic variables such as ground reaction forces [12, 16–19] and joint moments [13, 17–20]. In addition, a few studies investigated the use of IMU data in a neural network model to estimate the muscle activity of lower limb muscles during healthy subject’s walking [21, 22]. However, the previous studies proposed a neural network model to predict muscle activation patterns based on the raw IMU data (i.e., acceleration and angular velocity) that are not as straightforward as joint kinematics to further understand walking function. Due to the complexity of extracting meaningful information from the IMU data, the previous models may not provide a clear insight into the potential relationship between the input (i.e., IMU-based data) and the output (i.e., muscle activation patterns). Considering that well-established tools have been introduced to analyze joint kinematics during dynamic movements such as walking and running, a neural network model based on joint kinematics can enhance our understanding of the potential relationships between joint angles and muscle activation patterns, potentially allowing more intuitive interpretation of the outcomes and providing insights into diagnosis and treatment planning in real clinical settings.

The purpose of this study is to demonstrate a proof-of-concept for the feasibility of using a complete lower extremity joint angle data (i.e., three plane angles of pelvis, hip, knee, and ankle joints), collected from healthy adults who performed level walking trials at different speeds [23], to train a deep-learning model to predict muscle activation patterns of the soleus muscle. This research would contribute to a shift from a primary reliance on physician observation (qualitative) towards the addition of a more objective measure (quantitative) in clinical settings.

## Methods

### Dataset description

This study used an open dataset [24] to build a deep-learning model to predict the soleus muscle activation patterns from the 12 lower limb joint angles (i.e., three plane angles of pelvis, hip, knee, and ankle joints) during level walking at different speeds.

The original dataset is comprised of 50 healthy subjects (age: 27.0 ± 17.6 years; height: 160.1 ± 19.8 cm; body mass: 56.7 ± 19.8 kg, F/M: 25/25) without any known locomotor disorders or health complications that would impact motor performance. Out of the 50 subjects’ data, 20 subjects’ data (age: 39.7 ± 15.0 years; age range: 26–72 years; height: 168.7 ± 10.5 cm; height range: 154.0–187.0 cm; body mass: 67.0 ± 13.0 kg; body mass range: 50.0–102.0 kg; F/M: 11/9; walking speed: 1.2 ± 0.4 m/s; walking speed range: 0.4–2.1 m/s) were used for the model training, validation, and test process. Twenty-one subjects were excluded due to their age <18 years old, and nine subjects due to considerable deviation from the normative soleus activation patterns during walking [25] (e.g., excessive muscle activity during the swing phase, and highest peak value occurring too early or too late). Each subject performed a series of locomotion tasks at their natural speed with no detailed instruction about gait speed or cadence to maintain the subject’s typical gait and then performed another set of trials in which they successively increased or decreased their speed.

The original dataset includes the joint angles for the pelvis, hip, knee, and ankle, and the raw EMG data for the eight lower extremity muscles (tibialis anterior, soleus, gastrocnemius medialis, peroneus longus, rectus femoris, vastus medialis, biceps femoris, and gluteus maximus) from both legs during four movements: level ground walking, toe-walking, heel-walking, and stair walking (step ascending and step descending). The data were summarized as a complete stride (i.e., two consequent initial contacts of the same limb) [24]. Out of the available described data, only the joint angles and the raw soleus EMG data on the right leg during level ground walking at different speeds were used to demonstrate the feasibility in this proof-of-concept study. Additional detailed information on the experimental protocol is described in [24].

### Data preprocessing

Considering that the raw EMG data segmented according to a single gait cycle are likely distorted due to the transient response of a digital filter, two additional copies of a given segmented raw EMG data were added following the original data upon an assumption that normal walking is a cyclic motion (i.e., as if the EMG data were collected for three gait cycles). The EMG data were then band-pass filtered using a fourth-order Butterworth filter with a frequency band of 20–450 Hz. If the EMG data were collected at a sampling frequency below 1 kHz, they were high-passed filtered using a fourth-order Butterworth filter with a cutoff frequency of 20 Hz. Full-wave rectification was performed, followed by a moving average with a sliding window of 400 ms, normalization to the maximum value of the respective trial, and segmentation of the middle bounds of the expanded EMG data. The processed EMG data and the original joint angles were then downsampled to 120 Hz, rather than the constant data length, in order to preserve the original walking speed information. The data preprocessing described was conducted in MATLAB (R2023a, The MathWorks, Inc., Natick, MA, USA).

### Deep-learning model architecture

The proposed model architecture is shown in Figure 1. The input variables are 12 joint angles, and the prediction variable is a soleus muscle activation pattern over one gait cycle during level ground walking. To effectively handle both input and output time-series data, the model architecture was based on the long short-term memory (LSTM) and fully connected layers. A sequence input layer with 84 features (i.e., 6-point delay × 12 joint angles) was used, followed by an LSTM layer with 32 hidden units, a fully connected layer with an output size of 16, a dropout layer of 20%, another fully connected layer with an output size of 1, and a regression layer. The hyperparameter values were adjusted to find the optimal configuration following monitoring of the training process and reviewing two test metrics such as root mean square error (RMSE) and loss.

**Figure 1.**
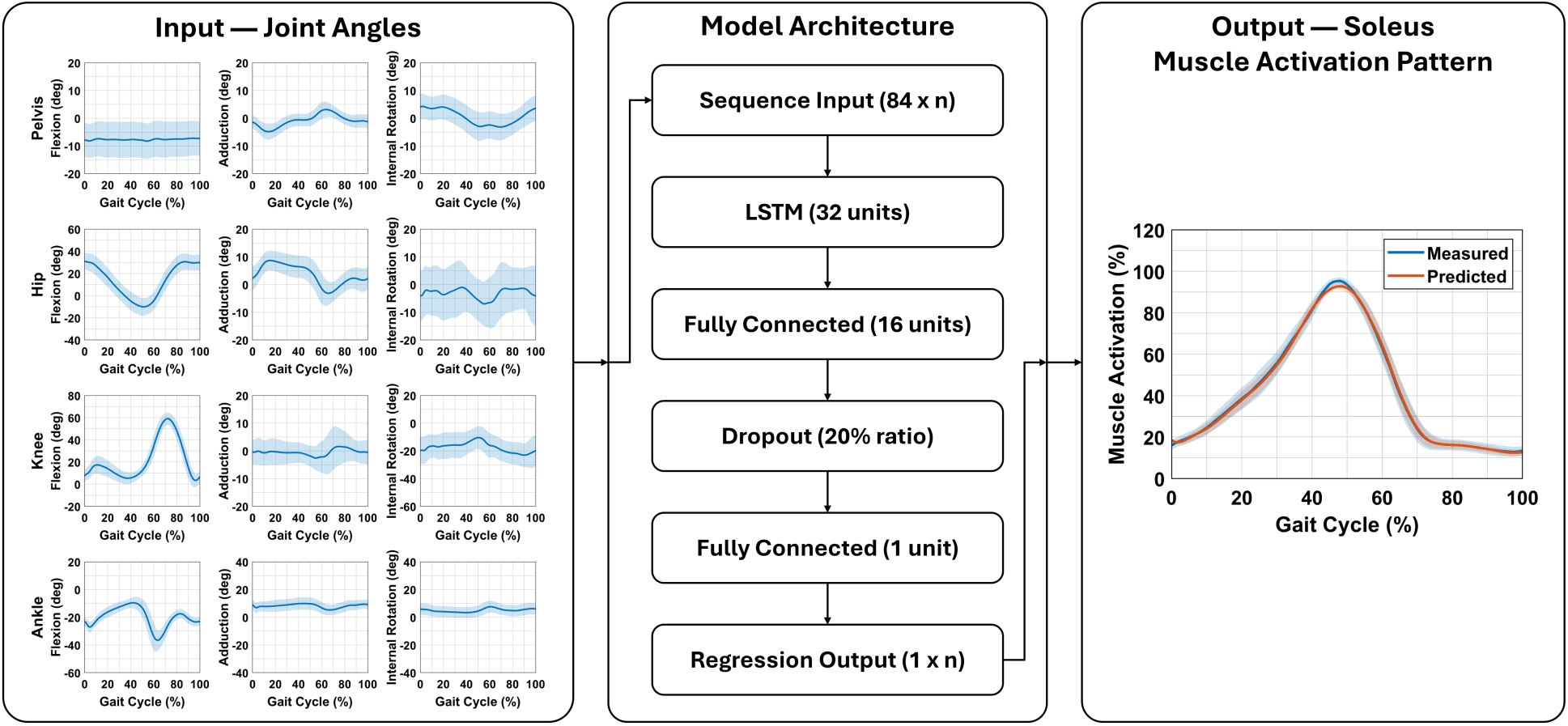
Architecture of the proposed deep-learning model that predicts soleus muscle activation patterns from twelve lower-extremity joint angles in three anatomical planes during normal level walking. Note that n indicates the number of time frames of input and output data.

### Model training and evaluation

The proposed model was implemented, trained, and tested in MATLAB (R2023a, The MathWorks, Inc., Natick, MA, USA). All layer arguments, but the regression layer, were defined as described in the Deep-Learning Model Architecture section. The regression layer used default arguments. The adaptive moment estimation (Adam) optimizer was used to minimize the mean squared error during the model training. A mini-batch size of 8, maximum number of epochs of 30, and epoch shuffle after every epoch were selected. An L2 regularization factor of 0.0001 was used. The initial learn rate was set to 0.00015, and the learn rate was dropped by a factor of 0.15 every epoch using the piecewise learning rate schedule. The training options were selected based on observation of the converging training and validation curves for RMSE and loss, and two acceptable performance metrics (i.e., RMSE and coefficient of determination, *R*^2^, of relationships between the measured and predicted muscle activation patterns). During the training process, the validation loss was calculated at the end of each epoch, and the model with the best validation loss out of 30 epochs was saved as the final model automatically.

The developed models were evaluated, using two different cross-validation approaches: (1) random data split cross-validation, called CVM1; and (2) leave-one-subject-out cross-validation, called CVM2. For CVM1, all the trials from all 20 subjects were split randomly into a training dataset (80%), validation dataset (10%), and test dataset (10%). For CVM2, the trials from all subjects except one were split randomly into a training dataset (80%) and validation dataset (20%), and the trials from the left-out subject was used as a test dataset, considering that the intended future application of the developed model is to predict the muscle activation patterns from a new patient. Since the outcomes may vary due to the random nature introduced in the data shuffling option, dropout layer, and random data split, the network training was repeated 10 times for the CVM1 and repeated 10 times for each subject model for the CVM2. Thus, there were 10 models of the CVM1 and 200 (i.e., 10 repetitions × 20 subjects) of the CVM2. For each model, we determined the RMSE, RMSE normalized to the range of the test data (NRMSE), and mean absolute error (MAE) between the measured and predicted muscle activation patterns, as the prediction performance metrics. Linear regression analyses were conducted to calculate additional performance metrics such as *R*^2^ and slope of relationships between the measured and predicted muscle activation patterns. Finally, the grand average and standard deviation values of the performance metrics from the 10 repetitions were used as the representative performance metrics for both cross-validation methods. For the error metrics (i.e., RMSE, NRMSE, and MAE), the smaller, the better. For the regression metrics (i.e., *R*^2^ and Slope), the closer to 1.0, the better.

In addition, further analyses were conducted to evaluate whether the models predict an accurate muscle activation timing (i.e., the difference between the measured and predicted times where the peak muscle activation was observed) and whether each performance metric is a function of age and walking speed, using a linear regression. The Mann-Whitney U test was used to test whether each performance metric is different between females and males. These analyses were also performed using MATLAB (R2023a, The MathWorks, Inc., Natick, MA, USA) with a significance level (α) of 0.05.

## Results

Figures 2 and 3 show a comparison between the measured and predicted soleus muscle activation patterns from the CVM1 and CVM2, respectively. The CVM1 predicted the muscle activation patterns well for the test dataset, as supported by both the small error metric values (i.e., RMSE: 0.075 ± 0.005; NRMSE: 0.086 ± 0.006; and MAE: 0.059 ± 0.004) and the regression metric values (i.e., *R*^2^: 0.948 ± 0.006; Slope: 0.956 ± 0.018; and *p* < 0.001). Similarly, the CVM2 also demonstrated a reasonable prediction performance with the relatively small error metric values (i.e., RMSE: 0.113 ± 0.016; NRMSE: 0.129 ± 0.023; and MAE: 0.091 ± 0.015) as well as the regression metric values (i.e., *R*^2^: 0.901 ± 0.071; Slope: 0.891 ± 0.033; and *p* < 0.001).

**Figure 2.**
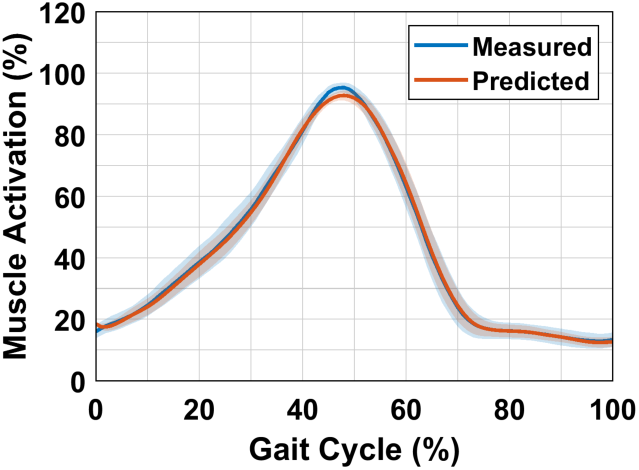
Model prediction of soleus muscle activation (predicted) compared to reference muscle activation (measured) for the random split cross-validation method (CVM1).

**Figure 3.**
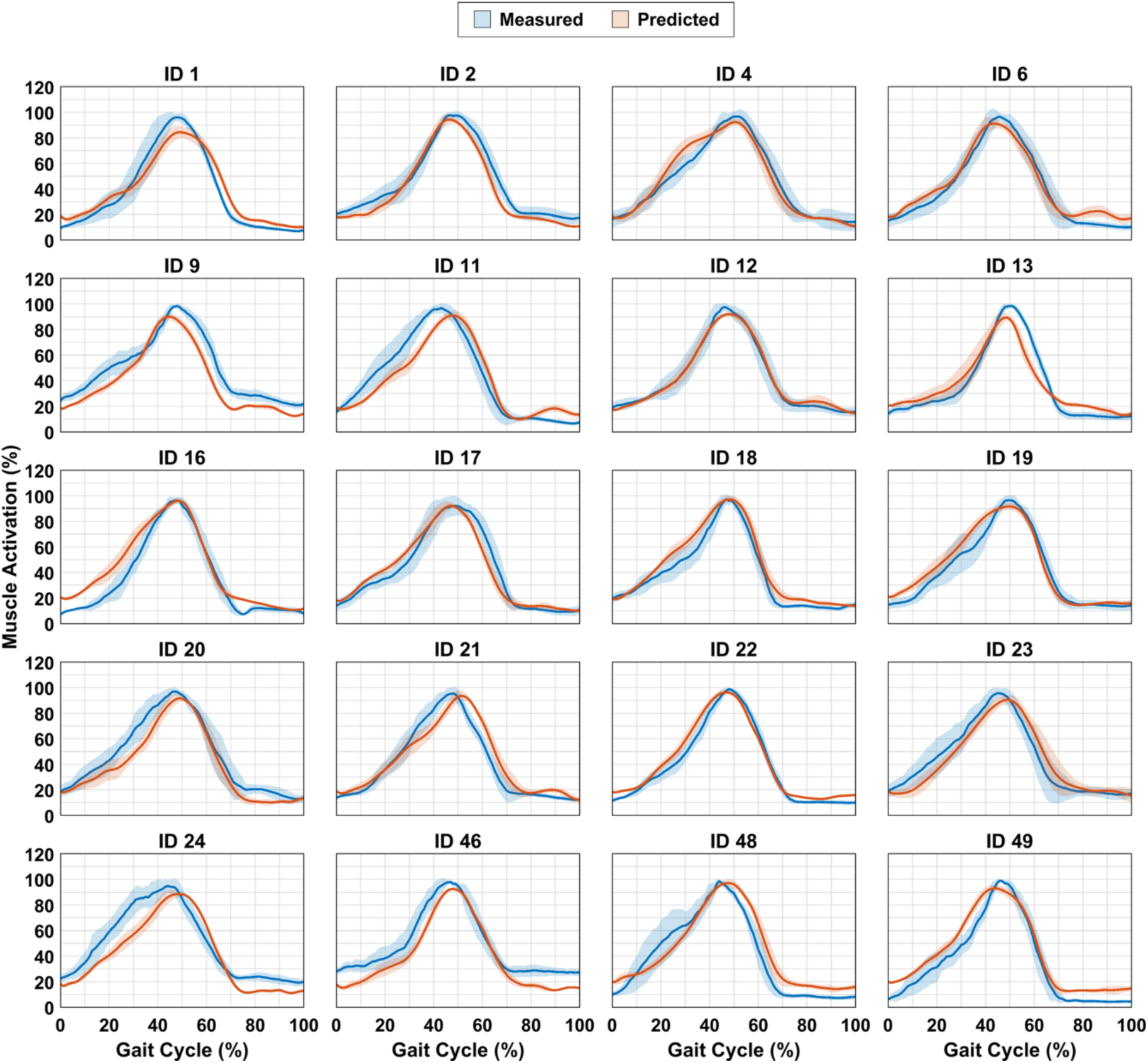
Model prediction of soleus muscle activation (predicted) compared to reference muscle activation (measured)for the leave-one-subject-out cross-validation method (CVM2). Each subplot represents a subject model with each title identifying with the corresponding subject ID from [23].

Figure 4 shows the prediction error of muscle activation timing for both cross-validation methods. On average, the errors from the CVM1 were positive for nine out of the ten repetitions, indicating that the CVM1 predicted the peak values ∼1% of the gait cycle later than the measured ones. The variability in the errors was also small, demonstrating that the prediction for most of the repetitions showed an average error of < 1%. However, the average error from the CVM2 was not consistent across the participants. The CVM2 yielded a small error (< ±2%) for 12 participants (i.e., ID 2, 4, 6, 12, 13, 16, 17, 18, 19, 22, 46, and 49) but a relatively larger error for the remaining eight participants. In addition, the peak values predicted with the CVM2 were found ∼3% earlier than the measured ones for eight participants (i.e., ID 2, 4, 6, 9, 13, 19, 22, and 49) but ∼7% later for the remaining 12 participants.

**Figure 4.**
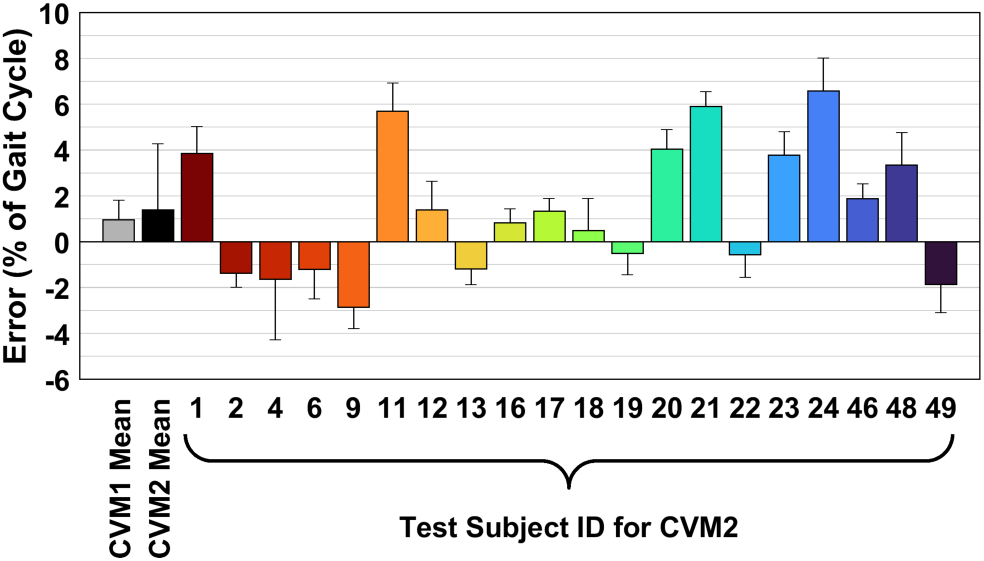
Average error of peak muscle activation timing for the random data split cross-validation (CVM1) and for the leave-one-subject-out cross-validation (CVM2). The numbered bars for the CVM2 represent the test subject ID from [23]. The error bars indicate the standard deviation of the ten repetitions.

The relationships between the performance metrics and age are shown in Figure 5. The CVM1 showed better performance metrics than the CVM2. However, regardless of the cross-validation models, the linear regression analyses clearly demonstrated that age has a negligible effect on the performance metrics, as supported by non-significant relationships for the CVM1 (*R*^2^ range: 0.003–0.193; and *p*-value range: 0.133–0.863) and the CVM2 (*R*^2^ range: 0.000–0.067; and *p*-value range: 0.270–0.933).

**Figure 5.**
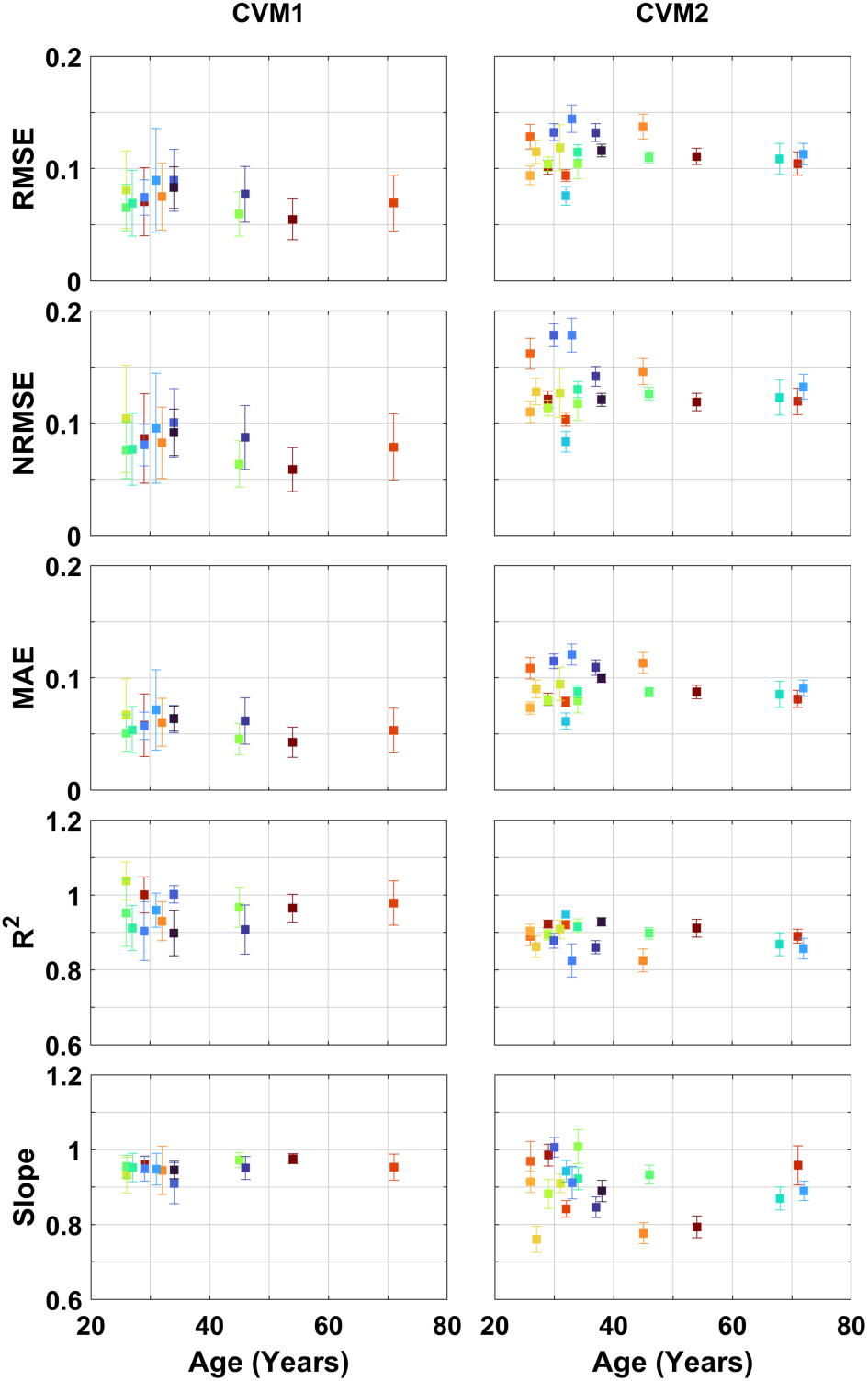
Scatterplots of relationship between the average performance metrics and age for the random data split cross-validation (CVM1) and for the leave-one-subject-out cross-validation (CVM2). Each color represents a different subject.

The performance metrics were not significantly different between female and male participants in most of the repetitions. For each of the performance metrics, the CVM1 yielded only one significant repetition out of ten (*p* < 0.029). However, there was no significant difference in all the performance metrics between the sexes using the CVM2 (*p*-value range: 0.058–1.000).

For the CVM1, walking speed was not significantly correlated with the performance metrics in most of the repetitions (Figure 6). The grand average of the *R*^2^ values of the relationship between the performance metrics and walking speed among ten repetitions was ∼0.1. Although the CVM2 generally showed low-to-moderate relationships with a few significant repetitions for most of the test subjects, moderate-to-high relationships were also predicted for some participants (e.g., ID 11, 13, 46, 48, and 49 for the RMSE, NRMSE, MAE, and *R*^2^; and ID 12, 20, and 49 for Slope).

**Figure 6.**
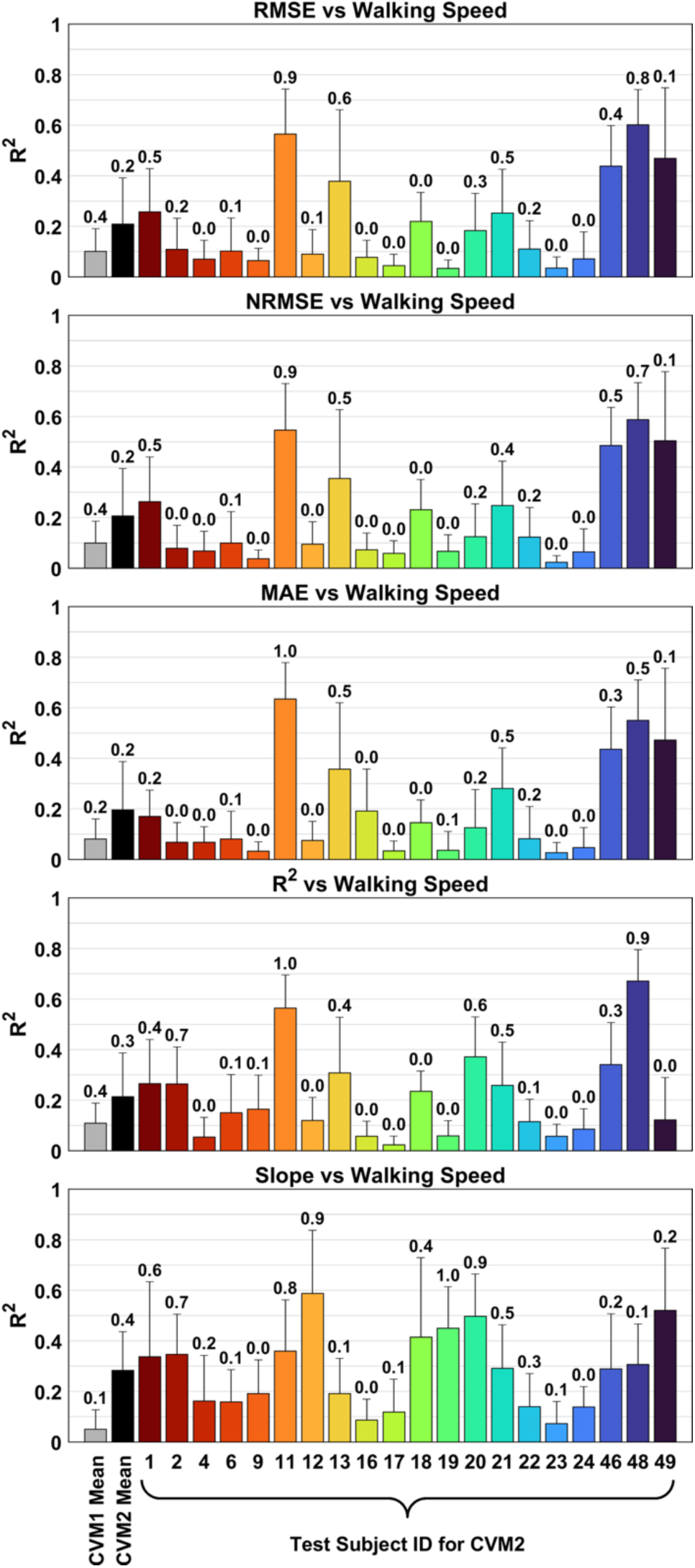
Summary of coefficients of determination (R2) of relationship between the average performance metrics and a subject’s walking speed in trial for the random data split cross-validation (CVM1) and for the leave-one-subject-out cross-validation (CVM2). The numbered bars for the CVM2 represent the test subject ID from [23]. The error bars indicate the standard deviation of ten repeated training trials, and the numbers for each error bar represent the percentage of significant repetitions out of the ten repetitions.

## Discussion

The objective of this study was to demonstrate the feasibility of using lower-limb joint angles for a deep-learning model to predict soleus muscle activation patterns collected from healthy adults during level walking at different speeds. The LSTM-based neural network models were developed to predict soleus muscle activation patterns and then evaluated using two different cross-validation methods (i.e., CVM1 and CVM2). Our main findings include that (1) the developed models performed reasonably well, as supported by the small error metric values as well as regression metric values close to 1.0 for both cross-validation methods; (2) the predicted muscle activation timing was reasonably close to the measured one when evaluating with the CVM1 (i.e., average error of < 1% of gait cycle), whereas the average error was affected by the new test dataset when evaluating with the CVM2 (i.e., grand average error of ±4% of gait cycle); and (3) the CVM1 demonstrated a negligible effect of age, sex, and walking speed on the performance metrics, while the performance metrics appeared to be influenced by the new test dataset when evaluating with the CVM2. Collectively, these results show the potential of the developed models to study muscle activation patterns using the lower-limb kinematics.

There have been attempts to estimate muscle activation patterns of several lower limb muscles from IMUs-based gait features [22; 23]. Khant et al. proposed an LSTM neural network based on four cascaded layers with the hidden units of 256 each, and the proposed models predicted the soleus muscle activation patterns with a correlation coefficient of 0.96 for both test and unseen data [22]. In their later study, they introduced feedforward neural networks with two extraction methods with a correlation coefficient of 0.56–0.96 for unseen data that predict muscle activation patterns of several lower limb muscles but not including the soleus muscle [23]. Although these previous studies showed promising results that muscle activation patterns could be predicted from wearable sensor data, these studies did not report a slope of the relationships between the measured and predicted muscle activation patterns and between the performance metrics and some potential factors (e.g., age, sex, and walking speed), leading to an uncertainty on whether the proposed models underpredict or overpredict the outcomes and whether the prediction performance can be a function of other factors. Moreover, as the previous models used angular velocities and accelerations as input data, understanding the relationship between the IMUs-based gait features and muscle activation patterns may not be straightforward especially for clinical applications.

In this study, we developed an LSTM-based deep-learning model to predict the soleus muscle activation patterns from lower limb joint kinematics, using a simpler model architecture (i.e., one LSTM layer with 32 hidden units and one dense layer with 16 units) than that of the previous study (i.e., four LSTM layers with 256 units each) [22]. The CVM1 and CVM2 yielded an average correlation coefficient of 0.96 and 0.83– 0.95, respectively, which is comparable to the previous studies. In addition, the prediction of the developed models appears not to be affected significantly by the potential factors such as age, sex, and walking speed, although the current samples skewed on the younger bound of the age range (26–72 years old) as only 30% of the sample was 40+ years old.

In addition, our proposed model may further provide an insight into the potential relationship between joint kinematics data (input) and muscle activation patterns (output), as supported in part by our preliminary analysis with the CVM1 (Figure 7) using the feature ablation method. Changes in the performance metrics with the removal of each joint contribution (i.e., zero flexion, abduction, and rotation angles of one of the four joints) were conducted to determine the relative importance of each joint to the prediction of muscle activation patterns. Although the removal of each joint did not change overall activation patterns as supported by the high *R*^2^ values (range: 0.797–0.917), some features of the patterns such as the peak values and the timing of the peak values seem to be affected by the hip joint angles (RMSE: 0.182 ± 0.018; NRMSE: 0.209 ± 0.020; MAE: 0.141 ± 0.012; and Slope: 0.590 ± 0.051), followed by the ankle (RMSE: 0.161 ± 0.017; NRMSE: 0.184± 0.019; MAE: 0.120 ± 0.010; and Slope: 0.673 ± 0.061), knee (RMSE: 0.138 ± 0.021; NRMSE: 0.158 ± 0.025; MAE: 0.106 ± 0.015; and Slope: 0.739 ± 0.059), and pelvis (RMSE: 0.097 ± 0.007; NRMSE: 0.112 ± 0.008; MAE: 0.077 ± 0.006; and Slope: 0.937 ± 0.031). Of the four joints, the removal of the pelvis joint angles had the least negative impact, suggesting its minimal contribution to model prediction performance. Whereas the hip joint angles showed to be critical as its removal significantly degraded model prediction performance, which might be counterintuitive, so that the further studies are needed. These preliminary findings demonstrated the potential that the developed model can potentially enhance our understanding of relationships between the altered joint angles and abnormal muscle activation patterns in future clinical applications.

**Figure 7.**
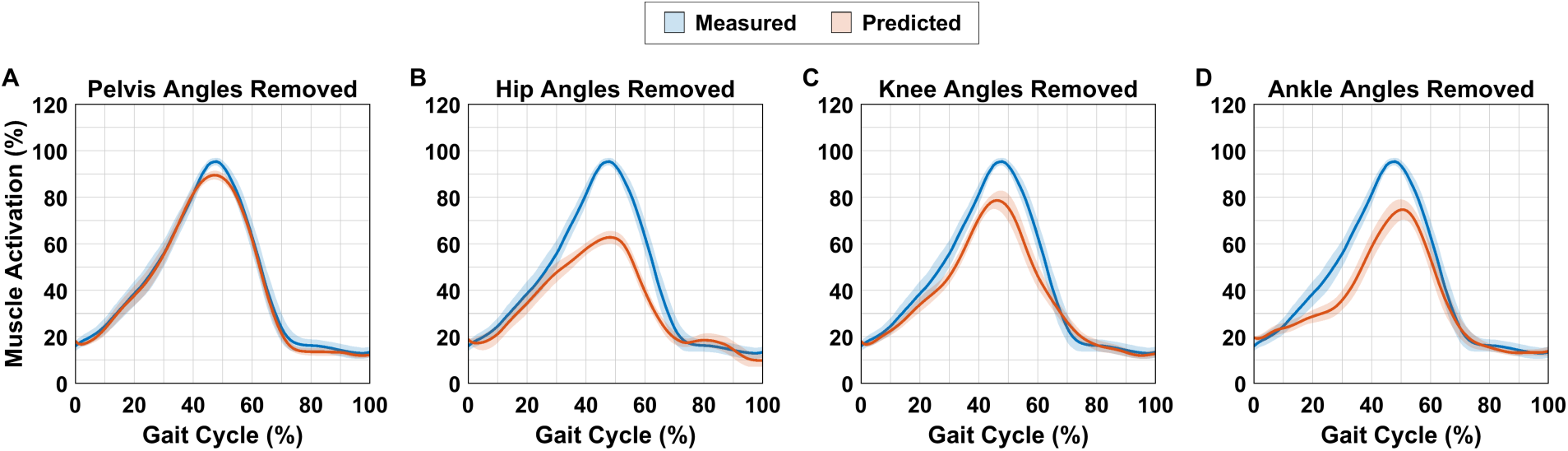
Preliminary results of relative contribution of lower limb joints to soleus muscle activation patterns using feature ablation method. Model prediction of soleus muscle activation (predicted) with the removal of pelvis (A), hip (B), knee (C), and ankle (D) joint angles compared to reference muscle activation (measured) for the random split cross-validation method (CVM1).

There are several limitations to consider. First, the developed models were trained and tested using the small dataset of the level walking of healthy adults, and thus the current findings would not be generalizable for other locomotion conditions and/or for other populations (e.g., an individual aged <18 years old or an individual with movement impairment). Second, as the joint angles computed from the 3D motion capture system were used, it is currently uncertain whether the proposed models would yield a comparable performance when using the joint angles determined from other modalities such as goniometers and IMUs as well as from other protocols. Lastly, the feasibility of the proposed models using the joint angles to predict muscle activation patterns was demonstrated only for soleus, so it would be required to test for other muscles.

## Conclusion

In this study, we developed deep learning models for predicting soleus muscle activation patterns from lower-limb joint angles during level ground walking at different speeds, and evaluated the prediction performance, using two different cross-validation methods. The main findings demonstrated the feasibility of the proposed models to be a template for a potential tool that quantifies muscle activation patterns from joint angles. Future studies are required to improve the proposed model for other muscles, various tasks, and diverse populations.

## Data Availability

All data produced in the present study are available upon reasonable request to the authors.

## Acknowledgments

We would like to express our sincere gratitude to Lencioni and the team for sharing the dataset.

